# Effect of COVID-19 during pregnancy: Studying the maternal and neonatal outcomes and assessing the placental changes related to SARS-CoV-2

**DOI:** 10.1101/2022.11.29.22282903

**Authors:** Surabhi Madan, Dharshni Ramar, Devang Patel, Amit Chitaliya, Nitesh Shah, Bhagyesh Shah, Vipul Thakkar, Hardik Shah, Rashmi Chovatiya, Pradip Dabhi, Minesh Patel, Amit Patel, Nirav Bapat, Parloop Bhatt, Aarya Naik, Manish Rana, Himanshu Nayak, Karun Dev Sharma, Prashant Parikh, Bhavna Mehta, Bhavini Shah

## Abstract

**Background:** Pregnant females affected with COVID-19 are reported to have poorer disease outcomes as compared to non-pregnant females of a similar age group. COVID-19 may lead to adverse changes in the placenta, which needs to be studied.

**Methods:** This is a case series of 63 pregnant women hospitalized with COVID-19 from May 2020 to February 2021.The primary outcomes were maternal death or complications.

**Results:** 63 women were studied. 83.3% of women were in the age group of 26 to 35 years. 33% women had associated comorbidities. 68.3% of women tested positive in their third trimester, 15.9% and 11% tested positive in their second and first trimesters respectively. 73% women had mild disease and 27% women required oxygen support. 3/63 women died. One woman in the second and two women in the third trimester died respectively. Histopathological examination in 13 placentae (of 19 placentae examined) were suggestive of maternal and fetal malperfusion.

**Conclusion:** Pregnant COVID-19 women may develop disease-related as well as obstetric complications.

## Introduction

The COVID-19 pandemic caused by the novel coronavirus SARS CoV-2 (1) has caused an unprecedented public health emergency all over the world. Though the overall rates of adverse neonatal outcomes like stillbirth and death are low in women with suspected or confirmed COVID-19, pregnant women with COVID-19 are at a higher risk of hospitalization, intensive care unit admission, need for mechanical ventilation, death, risk of preterm delivery, and their babies being admitted to the neonatal unit; as compared to pregnant women without COVID-19 (2, 3). There is also an increased risk of obstetric complications in these women in addition to maternal mortality (4). Physiologically predisposing factors such as elevation of the diaphragm, decreased residual lung functional capacity, increased oxygen intake, and immune modulation during pregnancy are some of the possible reasons (5). Though the overall rate of congenital infection has been reported to be low, the extent of vertical transmission (in utero, intrapartum, early postnatal period) remains unclear (6).

While there are documented changes to the placenta due to infections by viruses such as Cytomegalovirus, Rubella virus and Zika virus; the similar information in COVID-19 is evolving (7, 8, 9).

The objectives of our study were to describe the clinical profile and outcome of the females admitted with COVID-19 during pregnancy and in the immediate post-partum period, to observe the neonatal outcomes, and to describe the histopathological changes in the placenta.

## Methodology

### Study design and setting

This is a case series of 63 pregnant female women hospitalized with COVID-19 from May 2020 to February 2021 in a tertiary care private hospital in western India. The study was approved by the CIMS (Care Institute of Medical Sciences) hospital ethics committee (CTRI/2020/05/025247).

### Study Population and data collection

Women who were admitted during pregnancy or within 6 weeks post-partum with a positive SARS-CoV-2 RT-PCR within the study period were included in the study. Demographic details, pregnancy-related information, signs and symptoms, results of laboratory parameters, radiological investigations, ward and ICU progress notes, treatment and outcome details, and delivery details were obtained from the medical record files and the hospital’s intranet. The status of respiratory support on all the days during hospitalization was recorded. If the woman did not have any symptoms and was found to have a positive PCR report, she was considered to be asymptomatic. For women who provided informed consent, their placentae were collected for histopathological examination, and vaginal secretions and amniotic fluid were obtained for SARS-CoV-2 RT-PCR testing. Placental tissue from the pregnant females was collected, buffered in 10% formalin and sent for macroscopic and histopathological analysis. Placental changes were reported and noted in accordance with the Amsterdam Consensus Criteria (10). The nasal and oropharyngeal swabs of the newborns were sent for SARS-CoV-2 RT-PCR immediately after birth. Due to the absence of a neonatal isolation unit in our hospital during the initial part of the study period, neonates were transferred post-delivery to other hospitals from where the RTPCR data could not be procured. As a separate IgM test kit was not available, IgG against COVID-19 spike protein was tested for the neonates within 24 hours of birth, after obtaining consent from the parents.

### Treatment Protocol

COVID-19 positive women were admitted to our institute since the first week of May 2020. The treatment protocol was updated regularly and was based on the categorization of women into mild, moderate, and severe categories, as defined by the Government of India; wherein mild disease is defined as the presence of upper respiratory infection without evidence of hypoxia; moderate disease is the presence of clinical features suggestive of pneumonia with SpO2 less than 94% on room air; and severe disease is clinical signs of pneumonia with SpO2 less than 90% on room air (11). Use of Remdesivir (RDV) in pregnant females was done at the discretion of the consultant, as per the available evidence. Steroids and other immunomodulatory drugs were used only in women with moderate to severe disease.

The primary outcomes were maternal death or complications. Secondary outcomes were neonatal death, positive SARS-CoV-2 PCR in the newborn, preterm birth and other neonatal complications.

## Results

63 women who tested positive for SARS-CoV-2 during pregnancy or within 6 weeks of delivery were admitted during the study period. 4/5^th^ (50/63) of women were in age group of 26 to 35 years, 9(14%) women were above the age of 35 years and 4(6%) women were in the age group of 21 to 25 years. 33% of women had associated comorbidities with the most common comorbidities being diabetes mellitus, hypothyroidism and hypertension while one woman each had pulmonary arterial hypertension, thalassemia with severe anemia, and chronic pulmonary disease. 44.4 % of the pregnant females were primigravida (28/63), 38.1 % were multigravida (24/63), 19% had a history of previous miscarriage (12/63) and 11% had a history of conception through in vitro fertilization (IVF) (6/63)/intrauterine insemination (IUI) (1/63). 68.3 % (43/63) females tested positive in their third trimester, 15.9 % (10/63) and 11% (7/63) tested positive in their second and first trimesters respectively. 88% of females were symptomatic while 11% were asymptomatic at the time of diagnosis of COVID-19. The most common symptoms were fever (73%), cough (65%), fatigue (42%), bodyache (28%), dyspnea (21%), headache (19%), anosmia (8%), dysgeusia (6%), nausea (5%) and abdominal pain (3%).

73% of women had mild disease (46/63), 8% had moderate disease (5/63) and 19% had severe disease (12/63). 27% (17/63) of women required oxygen support. The highest oxygen support was O2 via NC in 7 women, NRBM in 2 women, HFNC in 1 woman, BiPAP in 2 women, MV in 3 women and ECMO in 2 women. Table 1 represents the different modalities of oxygen support in different age groups. The number of women who needed oxygen support was highest in the age group of 31-35 years (30%), among which 2 women required ECMO. 9/17 (52.9%) women who needed oxygen support had no comorbidities, which includes the 5 women who required MV and ECMO supports.

**Table 1:**
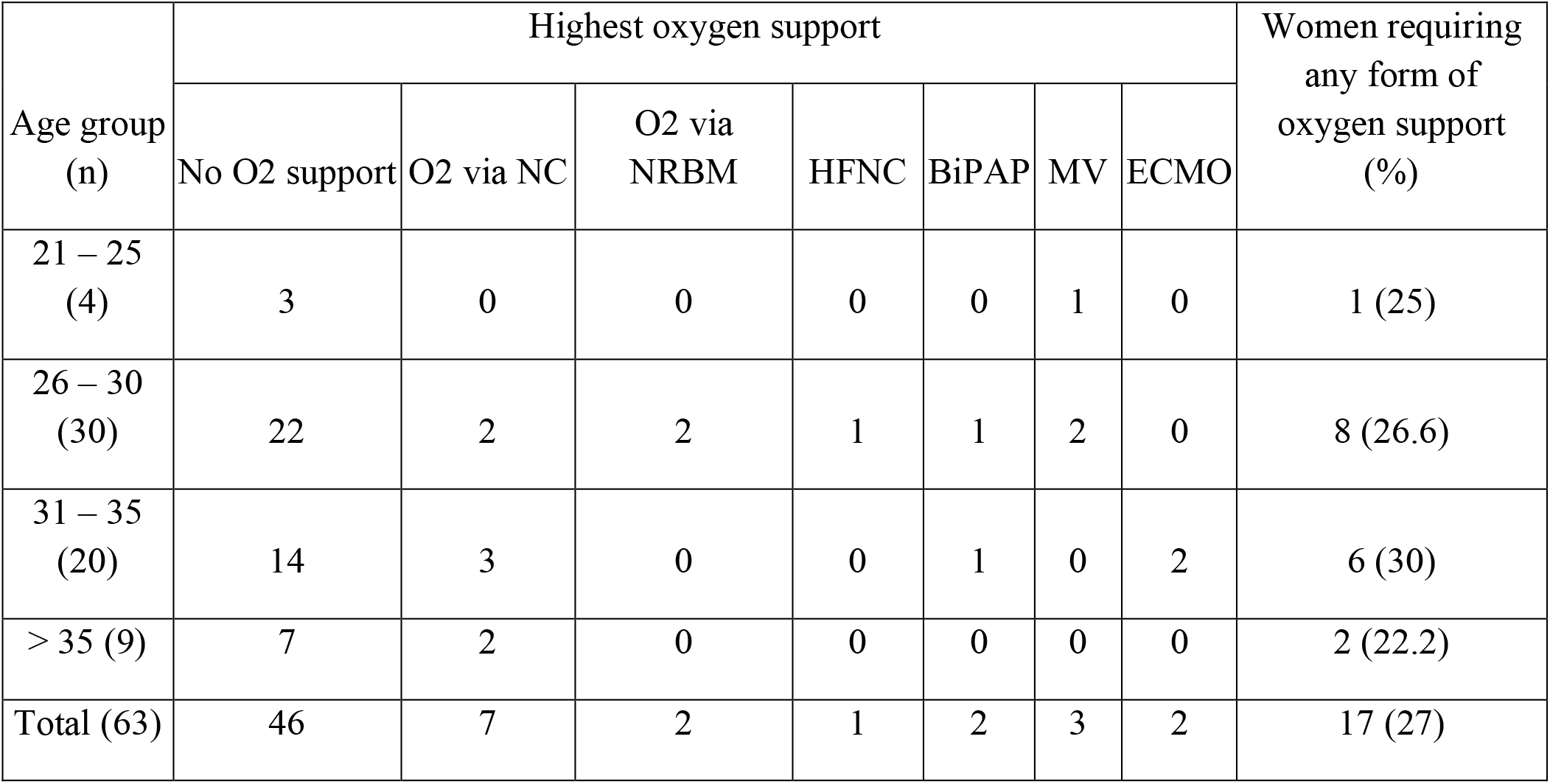
Oxygen requirement in different age groups.

Table 2 represents the oxygen requirement in different trimesters. No woman in the first trimester required oxygen support, as compared to 1/11 (9.1%) and 16/45 (35.6%) women who were diagnosed with COVID-19 in the second and third trimesters respectively. Similarly, there was no mortality in the first trimester, while one woman in the second and two women in the third trimester died respectively.

**Table 2:** Oxygen requirement in different trimesters.

| Trimester | Highest oxygen support |  |  |  |  |  |  |
| --- | --- | --- | --- | --- | --- | --- | --- |
|  | No support<br>n (%) | O2 via NC<br>n (%) | O2 via NRBM<br>n (%) | HFNC<br>n (%) | BiPAP<br>n (%) | MV<br>n (%) | ECMO<br>n (%) |
| 1st Trimester (n =7) | 7 (100) | 0 (0) | 0 (0) | 0 (0) | 0 (0) | 0 (0) | 0 (0) |
| 2nd Trimester (n=11) | 10 (90.9) | 0 (0) | 0 (0) | 0 (0) | 0 (0) | 0 (0) | 1 (9.1) |
| 3rd Trimester (n=45) | 29 (64.4) | 7 (15.6) | 2 (4.4) | 1 (2.2) | 2 (4.4) | 3 (6.8) | 1 (2.2) |
| Total | 46 | 7 | 2 | 1 | 2 | 3 | 2 |

Blood, lower respiratory tract secretions, abdominal secretions and urine samples were sent for cultures in 13/63 (20.6%) women, where secondary bacterial or fungal infections were suspected. Positive cultures were obtained in 6 women. Organisms isolated from blood were *Klebsiella pneumoniae, Staphylococcus aureus* and *Stenotrophomonas maltophilia*, while the respiratory samples were positive for *Stenotrophomonas maltophilia* and *Acinetobacter*. One woman had urine culture positive for *Providencia rettgeri*.

29/63 (46%) women received RDV and an equal number received steroids. 7 (11 %) women were administered tocilizumab. 1 woman each received infliximab and tofacitinib. 22 women received both RDV and steroids.

Table 3 represents the various maternal complications. Preterm delivery, antepartum haemorrhage and pre-eclampsia were the commonest.

**Table 3:** Maternal complications observed while hospitalization.

| **Complication** | **n (%)** | **Complication** | **n (%)** |
| --- | --- | --- | --- |
| Pre term delivery | 7 (11.1) | Pneumothorax | 1 (1.6) |
| Antepartum haemorrhage | 6 (9.5) | Hepatic Injury | 1 (1.6) |
| Pre- eclampsia | 5 (7.9) | Cardiac Complications | 1 (1.6) |
| Eclampsia | 2 (3.2) | Multiorgan dysfunction | 1 (1.6) |
| Miscarriage | 2 (3.2) | Acute Kidney Injury | 1 (1.6) |
| Shock | 2 (3.2) | Ectopic Pregnancy | 1 (1.6) |
| Sepsis | 2 (3.2) |  |  |

35/63 (55.6%) women delivered during the hospitalization period. Vaginal delivery was performed in one woman while lower segment caesarean section (LSCS) was performed in 31/35 (88.6%) women. Laparotomy was performed in one female due to a ruptured ectopic pregnancy with hemoperitoneum, while 2 women underwent a dilation and evacuation procedure due to miscarriage and per vaginal bleeding respectively. There were 25 live births, 5 stillbirths and 2 intrauterine fetal deaths (IUFD). Neonatal outcome in different maternal age groups is shown in table 4. Of the 25 neonates, 17 (68%) did not have any complications, 7 (28%) were born with low birth weight and one was born with meconium-stained liquor. No newborn had any respiratory complications.

**Table 4:** Neonatal outcomes in different maternal age groups.

| Age group (years) | Live birth | IUFD | Still birth | D&E | Laparotomy for ruptured ectopic | Total |
| --- | --- | --- | --- | --- | --- | --- |
| 21 – 25 | 1 | 0 | 3 | 1 | 0 | 4 |
| 26 – 30 | 14 | 0 | 0 | 1 | 0 | 15 |
| 31 – 35 | 7 | 2 | 1 | 0 | 1 | 11 |
| > 35 | 3 | 0 | 2 | 0 | 0 | 5 |
| Total | 25 | 2 | 6 | 2 | 1 | 35 |

Note: One 24 years female woman gave birth to 2 still born neonates

17 vaginal and 15 amniotic fluid samples were collected for RT-PCR. Among these, only 1 vaginal swab was positive for SARS-CoV-2 and the rest were negative. The neonatal outcome in the woman with positive RT-PCR from the vaginal swab was IUFD. RT-PCR for SARS-CoV-2 was sent in 19 neonates. None of the neonates had a positive result. IgG antibody against the spike protein of the SARS CoV-2 was found to be negative in all the neonates.

Histopathological examination of placental samples was performed in 19 women. Abnormal changes suggestive of malperfusion or ischemia were found in 13 samples. (Table 5, Fig 1).

**Table 5:**
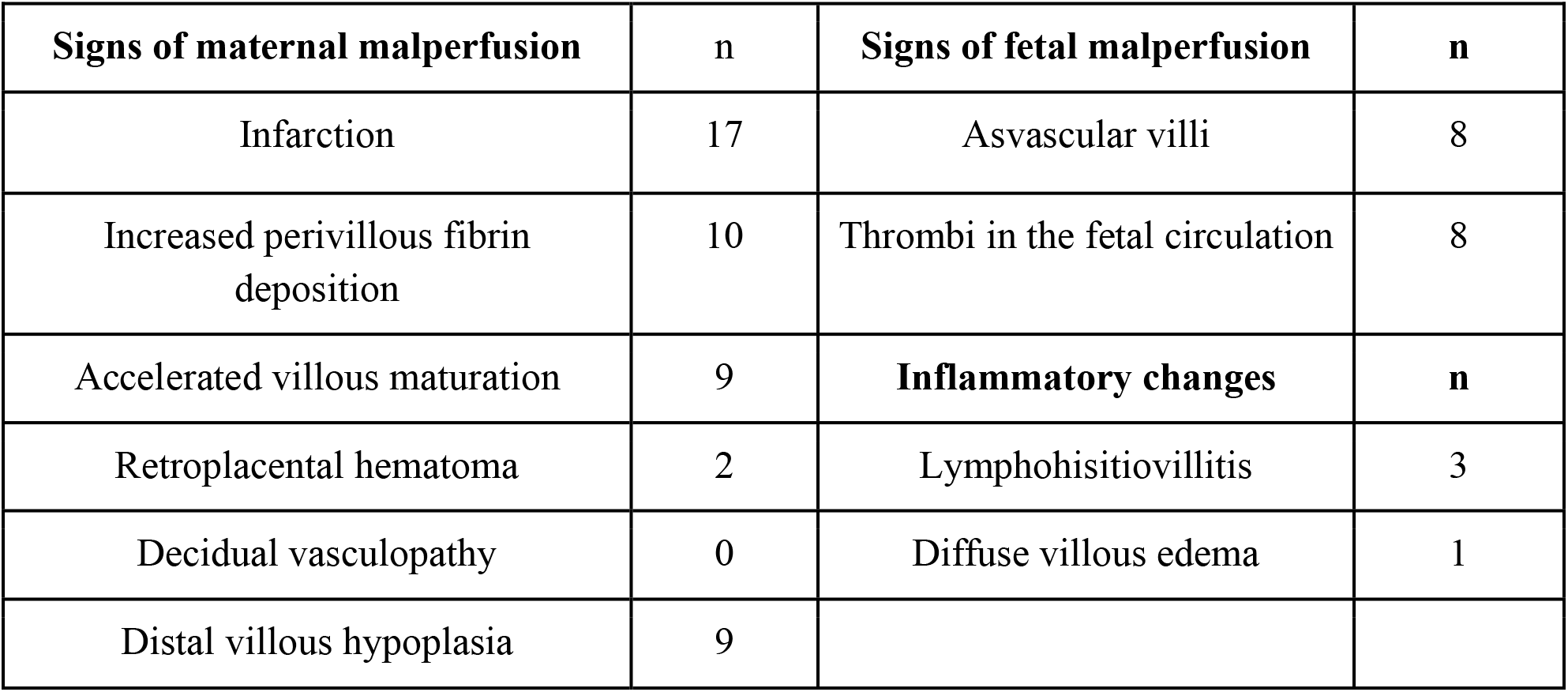
Placental findings.

**Fig. 1:**
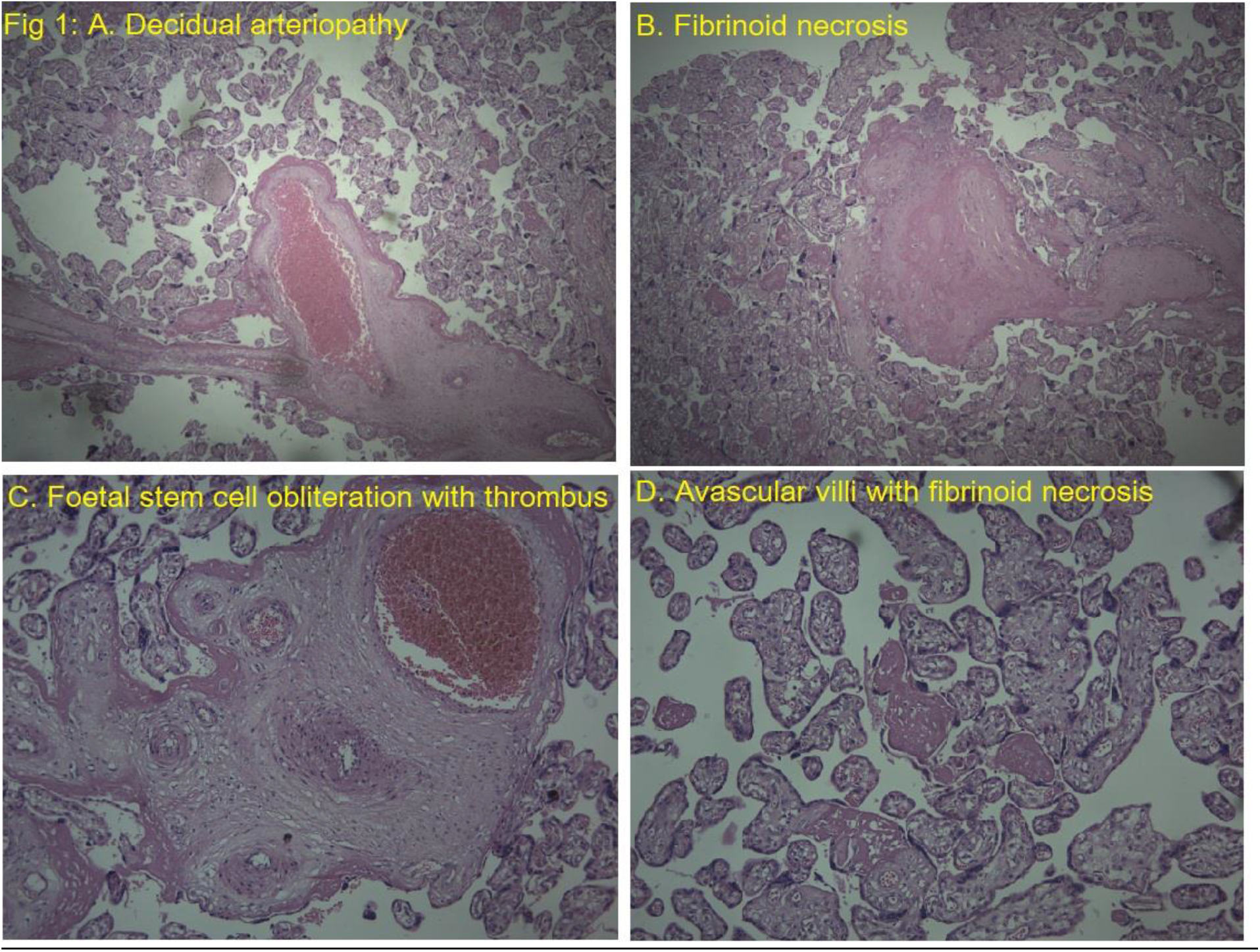
Histopathological changes seen in the Placenta.

Three (4.8%) women died; all these women had developed acute respiratory distress syndrome secondary to COVID-19 pneumonia. Two of them were in the age group of 30-35 years and one was in the age group of 20-25 years. Blood cultures of 2 of the women who died were positive *for Klebsiella pneumoniae* and *Staphylococcus aureus* and the lower respiratory culture of one of these two women was positive for *Acinetobacter*.

## Discussion

The present study highlights the effects of COVID-19 on maternal and neonatal outcomes. The study of histopathology of placentae of COVID-19 positive pregnant females was done to improve the knowledge and understanding of the placental changes in COVID-19 positive pregnant females.

In our study, 88% of women were symptomatic. Symptoms and signs of COVID-19 during pregnancy appear to be similar to those in non-pregnant individuals (2, 12). We observed maternal complications like antepartum haemorrhage, pre-eclampsia, eclampsia, preterm delivery, ectopic pregnancy, miscarriage and sepsis. In a large retrospective cohort study, SARS-CoV-2 infection was found to be associated with an increased risk for a composite outcome of maternal mortality or serious morbidity from the obstetric complications like hypertensive disorders of pregnancy, postpartum haemorrhage, or infection other than SARS-CoV-2; including a trend towards an increase in caesarean birth (4).

Data from the Centers for Disease Control and Prevention (CDC) has shown an increased risk of mechanical ventilation, ICU admission and death due to COVID-19 infection in pregnancy as well as an increased risk of preterm delivery and stillborn delivery (2, 3). In our study, 8% of the women had a moderate disease while 19% of women had severe disease. In various studies, risk factors for severe disease and death in pregnancy include older age (especially ≥35 years), obesity, preexisting medical comorbidities (particularly hypertension, diabetes, or more than one comorbidity), and being unvaccinated (13-15). No woman in our study was vaccinated, as recommendations for COVID-19 vaccination in pregnancy were not implemented in our country during the study period. In this study, the number of women who needed oxygen support and who died was highest in the age group of 31-35 years. Above 50% of women who needed oxygen support had no comorbidities.

Maternal infection after 20 weeks of gestation increased the risk of adverse obstetric outcomes, and maternal infection after 26 weeks increased the risk of adverse neonatal outcomes (16). In our study, oxygen requirement was the highest in the women who presented in the third trimester of pregnancy, and no woman in the first trimester needed oxygen support. While there was no mortality in the first trimester, one woman in the second and two women in the third trimester died respectively.

11% of the women in this study had preterm delivery. The risk of preterm and caesarian delivery has been observed mainly with a severe or critical disease in cohort studies (17, 18); underlying comorbidities also likely play a role. About 8% of women in this study had preeclampsia. A meta-analysis of observational studies of SARS-CoV-2 infection during pregnancy found 62 % higher odds of developing preeclampsia among women with COVID-19 (19). 6 stillbirths were noted in our study. Emerging data suggest an association between COVID-19 in pregnancy and stillbirth (3, 20, 21).

In our study, the nasopharyngeal swabs for SARS CoV-2 PCR of all the neonates were negative. These findings indicate a low risk of vertical transmission of the COVID-19 disease. This is identical to a study done in New York (22), which also concluded no evidence of vertical transmission of COVID-19 from the mothers to the neonates despite rooming-in and breastfeeding practices. IgG to SARS-CoV-2 was negative in all the neonates. The maternal IgG levels are highest more than 30 days after onset of symptoms (23). Factors affecting transplacental antibody transfer have been studied but are not clearly known (23,24, 25).

The overall neonatal outcome was good in this study. An analysis of data from pregnant women with confirmed or suspected SARS-CoV-2 infection in 12 countries reported all-cause early neonatal death rates of 0.2 to 0.3 percent (26). A systematic review also found that the incidence of neonatal death was similar among individuals who tested positive compared with negative for SARS-CoV-2 (27).

There are no standard criteria for diagnosis of placental SARS-CoV-2 infection and no definite COVID-19-specific placental changes (28). SARS-CoV-2 placentitis is characterized by chronic histiocytic intervillositis, increased perivillous fibrin deposition and villous trophoblast necrosis. These changes can cause widespread and severe placental destruction, resulting in placental malperfusion and insufficiency and leading to perinatal death from fetal hypoxic-ischemic injury. In a case series of 64 stillborns (15 to 39 weeks of gestation) and four neonatal deaths with SARS-CoV-2 placentitis, all 68 placentae tested positive for SARS-CoV-2, whereas the virus was detected from a stillborn/newborn body specimen in only 16 out of 28 cases tested (59 percent) (29). There was no evidence that fetal SARS-CoV-2 infection had a direct role in causing the deaths.

We observed placental changes of maternal underperfusion such as villous infarctions, increased perivillous fibrin deposition, accelerated villous maturation, distal villous hypoplasia and retroplacental hematoma. Fetal Vascular Malperfusion changes such as avascular villi and thrombi in fetal circulation were also noted (Fig 1). Moreover, inflammatory changes such as lymphohistiovillitis and diffuse villous edema were seen.

### Limitations

This study has the limitations of secondary data analysis. The sample size is small and there is no comparator arm. Also, the histopathological examination of the placenta and SARS-CoV-2 PCR could not be performed from the vaginal and amniotic fluid in all the women, which limited the information further.

## Conclusion

SARS CoV-2 infection during pregnancy and in the immediate post-partum period may be associated with adverse outcomes. The risk of disease severity and death may increase with the progression of pregnancy.

## Data Availability

All data produced in the present study are available upon reasonable request to the authors

## Informed Consent

A written informed consent for the use of patient information and images to be published was taken from each patient.

## Ethical Approval

The study was approved by the CIMS (Care Institute of Medical Sciences) hospital ethics committee (CTRI/2020/05/025247).

## Contributorship

Surabhi Madan, Devang Patel and Dharshni Ramar conceived the study and wrote the manuscript. Nitesh Shah, Bhagyesh Shah, Vipul Thakkar and Hardik Shah researched literature. Manish Rana and Nirav Bapat performed the analysis. Parloop Bhatt, Karun Dev Sharma, Himanshu Nayak and Mitesh Nayak approved the final manuscript. Surabhi Madan, Rashmi Chovatiya, Pradip Dabhi, Minesh Patel, Amit Patel, Arya Naik, Devang Patel collected and contributed data. All authors read and approved the final manuscript.

## Acknowledgements

The authors thank Dr. Riddhi Parekh, Dr. Disha Patel, Dr. Aditya Vyas and Aditi Naik; Department of Clinical Research, Care Institute of Medical Sciences, Ahmedabad, India, for their assistance in data aggregation and compilation in electronic database.

